# ALTERED MOLECULAR PATHWAYS OBSERVED IN NASO-OROPHARYNGEAL SAMPLES OF SARS-CoV-2 PATIENTS

**DOI:** 10.1101/2020.05.14.20102558

**Authors:** Emel Akgun, Mete Bora Tuzuner, Betul Sahin, Meltem Kilercik, Canan Kulah, Hacer Nur Cakiroglu, Mustafa Serteser, Ibrahim Unsal, Ahmet Tarik Baykal

## Abstract

**Background:** COVID-19 or severe acute respiratory syndrome coronavirus 2 (SARS-CoV-2) appeared throughout the World and currently affected more than 3.6 million people and caused the death of around 252,000 people. The novel strain of the coronavirus disease is transmittable at a devastating rate with a high rate of severe hospitalization even more so for the elderly population. Currently around 50,000 patients are in a seriously critical situation. Although 1.2 million patients recovered from the disease there are still more than 2.1 Million active cases. Naso-oro-pharyngeal swab samples as the first step towards detecting suspected infection of SARS-CoV-2 provides a non-invasive method for PCR testing at a high confidence rate. Furthermore, proteomics analysis of PCR positive and negative nasooropharyngeal samples provides information on the molecular level which highlights disease pathology.

**Method:** Samples from 15 PCR positive cases and 15 PCR negative cases were analyzed with nanoLC-MS/MS to identify the differentially expressed proteins.

**Results:** Proteomic analyses identified 207 proteins across the sample set and 17 of them were statistically significant. Protein-protein interaction analyses emphasized pathways like Neutrophil degranulation, Innate Immune System, Antimicrobial Peptides.

**Conclusion:** Neutrophil Elastase (ELANE), Azurocidin (AZU1), Myeloperoxidase (MPO), Myeloblastin (PRTN3), Cathepsin G (CTSG) and Transcobalamine-1 (TCN1) were found to be significantly altered in naso-oropharyngeal samples of SARS-CoV-2 patients. The identified proteins are linked to alteration in the innate immune system specifically via neutrophil degranulation and NETosis.

## 1. Background

Severe acute respiratory distress syndrome-associated coronavirus-2 (SARS-CoV-2) also known as COVID-19 first appeared in Wuhan, Hubei, China in December 2019, and now it is widespread around the World. The replication of the virus is through the human airway epithelial cells where it targets the receptors of human Angiotensin-Converting Enzyme 2 (ACE-2). The high mortality observed in COVID-19 is associated with severe acute respiratory distress and systemic coagulopathy. Many COVID-19 patients show a postponed onset of respiratory issues but then develop into more severe situations. We wanted to investigate the molecular changes in the COVID-19 patients’ naso-oropharyngeal swab samples via comparison to the proteome of PCR negative cases. Our goal is to find pathways associated with the site of infection through proteomics analysis. 17 statistically significant protein alterations lead us specifically to neutrophil degranulation pathways.

During airway infections, Neutrophils are the first wave of defense that also defines the disease outcome. Neutrophils have multiple functions in viral infections such as inactivation of the virus, achieved by phagocytosis, ROS production, proteolytic enzyme release, NET activation (NETosis, Neutrophil Extracellular Traps). Neutrophils also interact with immune cells and secreting cytokines participate in eliciting an antiviral response.

Neutrophil granulocytes express several enzymes linked to controlling host infection. As the cytotoxic molecules are released from the granules they have an impact on the inflammatory response. Such adverse molecules cause severe damage to the host where it exhibits itself as perivascular infiltrates around the capillaries in the lungs as observed in SARS-CoV-2 patients. The identified up-regulated proteins Neutrophil Elastase, Azurocidin, Myeloperoxidase, Myeloblastin, Cathepsin G, and Transcobalamine-1 (TCN1, AZU1, CTSG, PRTN3, MPO, ELANE) in naso-oropharyngeal swab samples are discussed to highlight the molecular mechanism changes in the site of infection.

## 2. Study design

### 2.1. Study Population and Sample Collection

Samples were collected using a nasal and oropharyngeal (NUCLISWAB) swab (Salubris, Turkey) and were placed in a tube with Universal Transport Media. After collection the samples were stored at –80 °C before proteomics analysis. The study group consists of 15 PCR positive and 15 negative cases. Informed consent was sought from each accepted to enter the study and ethical approval for the conduct of the study was given by Acibadem Mehmet Ali Aydinlar University Human Scientific and Ethical Review Committee (Approval ID: 2020–07/9).

### 2.2. COVID-19 RT-PCR Test

For molecular testing of SARS-CoV-2;

The extraction of nucleic acids from the samples was performed by a manual liquid phase method using Bio-Speedy Nucleic Acid Isolation Kit (Bioeksen, Turkey). Nucleic acid amplification test (NAT) was carried out by Bio-Speedy COVID-19 RT-qPCR Detection Kit(Bioeksen, Turkey) according to the manufacturer’s instructions on RotorGene (Qiagen, Germany) Real-Time PCR instrument. The test briefly achieves a one-step reverse transcription (RT) and real-time PCR (qPCR) targeting SARS-CoV-2 specific RdRp (RNA-dependent RNA polymerase) gene fragment

### 2.3. LC-MSMS Analysis

A shorter sample preparation approach was applied to obtain the tryptic peptides for analyses. 50 ul of the naso-oropharyngeal transport solution was taken and lyophilized. The powder was reconstituted in 20ul of 50 mM Ammonium bicarbonate solution with 1 ul of protease inhibitor mixture (Thermo Scientific). The mixture was centrifuged at 14000 xg for 10 min and the supernatant was transferred to a clean Eppendorf tube. DTT was added to 10 mM final concentration and incubated at 60°C for 10 min. The mixture was alkylated in dark for 30 min with 20mM IAA. To the resulting mixture 1 ug of sequencing grade trypsin (Promega Gold) was added and incubated at 55°C for 1.5 hrs. The digest was acidified with 1 ul formic acid and transferred to an LC vial for injection. The samples were analyzed by the protocols in our previous studies [1]. Briefly, tryptic peptides were trapped on a Symmetry C18 (5µm,180µm i.d. × 20 mm) column and eluted with ACN gradient (4% to 40% ACN, 0.3 ul/min flow rate) with a total run time of 60 min on a CSH C18 (1.7 µm, 75 µm i.d. × 250 mm) analytical nano column. Data were collected in positive ion sensitivity mode using a novel data-independent acquisition mode (DIA) coined as SONAR [2] with a quadrupole transmission width of 24 Da. Progenesis-QIP (V.2.4 Waters) was used for data processing.

### 2.4. PPI Network and Pathway Analysis

Protein-protein interaction networks functional enrichment analysis was carried out using the STRING (http://string-db.org/, v11.0) database with the highest confidence interaction score level to identify possible pathways related to the identified proteins. Textmining, experiments, databases co-expression, neighborhood, gene fusion, and co-occurrence selected as active interaction sources. The minimum interaction score was set to high (high confidence=0.700). REACTOME (http://www.reactome.org) pathways analysis tool was also used for processing.

## 3. Results

### 3.1. Label-free Proteomics

Proteins were extracted from a small amount of naso-oropharyngeal samples, fast tryptic digestion was applied and followed with a 60 min reverse-phase separation. NanoLC-MSMS analysis provided the identification of 207 protein groups with high confidence (<1% FDR). Statistical analysis done in Progenesis QIP software identified 17 proteins to be statistically significantly expressed in patients’ naso-oropharyngeal samples (Table 1).

**Table 1:**
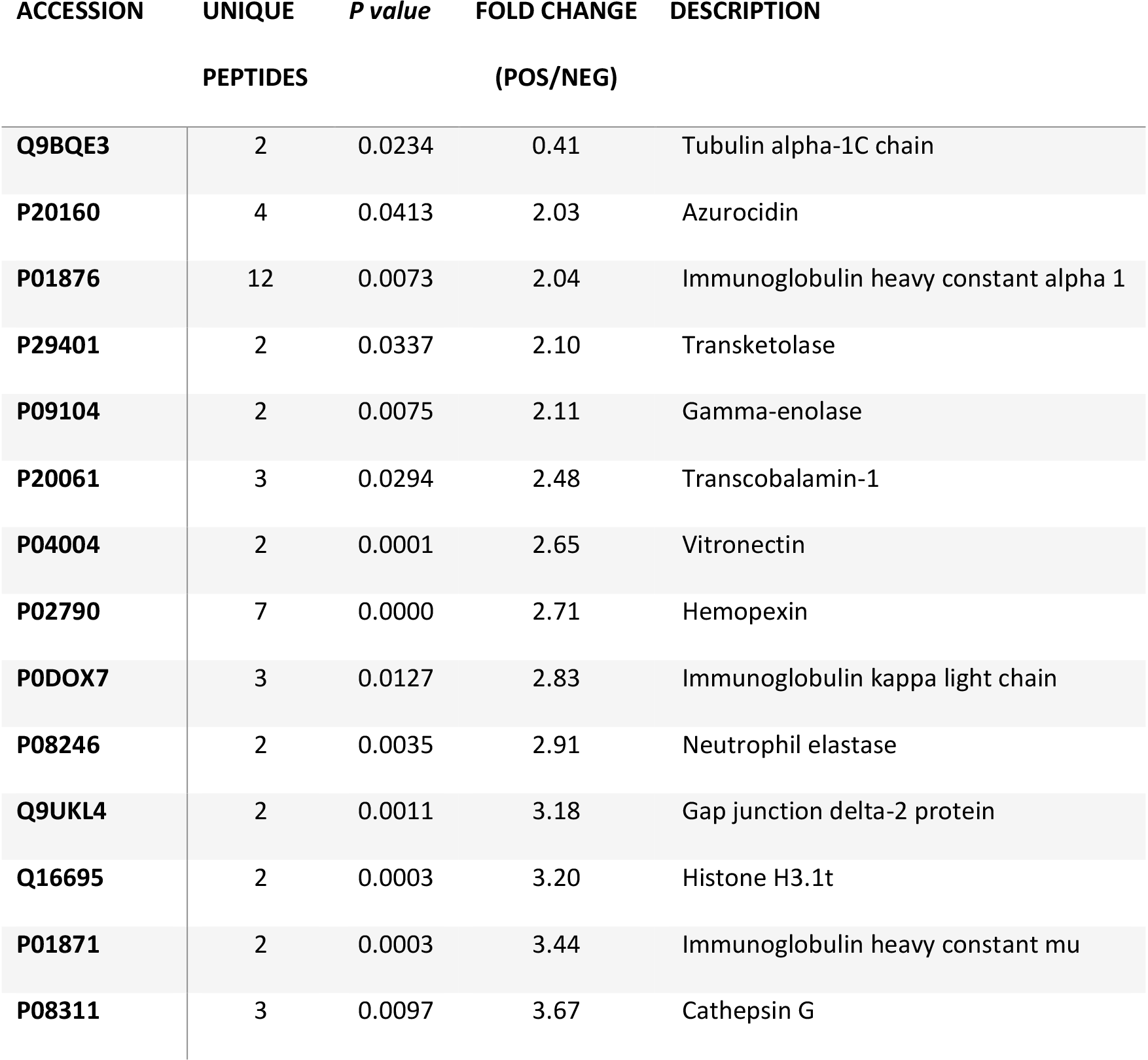
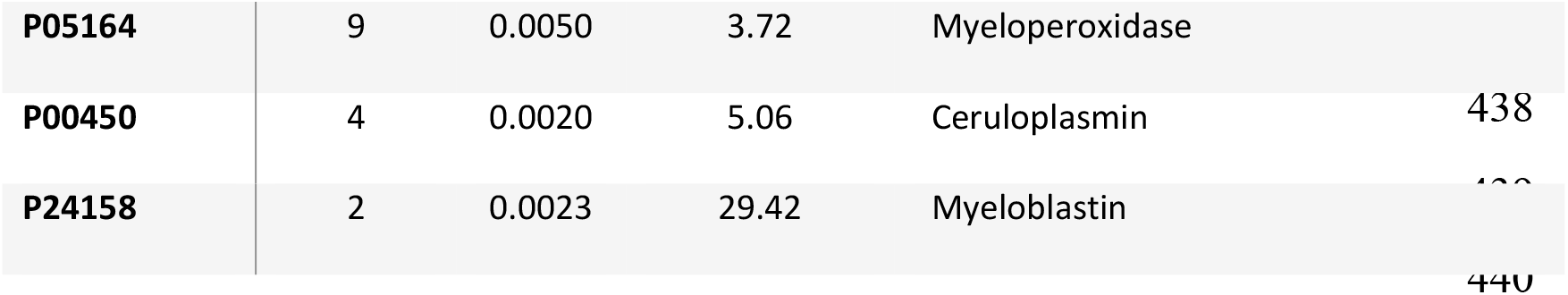
Significanltly altered protein identification list.

### 3.2. Bioinformatic Analysis

Pathway analysis of the significantly altered protein levels between COVID-19 positive and negative patients’ naso-oropharyngeal swab samples were analyzed using the STRING online database. The PPI network obtained, contained 17 differentially expressed proteins with 15 nodes (disconnected nodes were not shown) and 14 edges as shown in Figure 1A. The main cluster includes Neutrophil Elastase (ELANE), Azurocidin (AZU1), Myeloperoxidase (MPO), Myeloblastin (PRTN3), Cathepsin G (CTSG) and Transcobalamine-1 (TCN1). The abundance of these proteins were found to be increased in COVID-19 positive patient samples compared to negative ones. REACTOME pathway enrichment analyses of the differentially expressed proteins were performed. Nine of the proteins were primarily associated with the immune system pathway. Proteins clustered in the PPI network were mainly enriched in the neutrophile degranulation pathway (HSA-6798695, FDR=2.01E-7) which is a subpathway of the innate immune system (Figure 1B).

**Figure 1.**
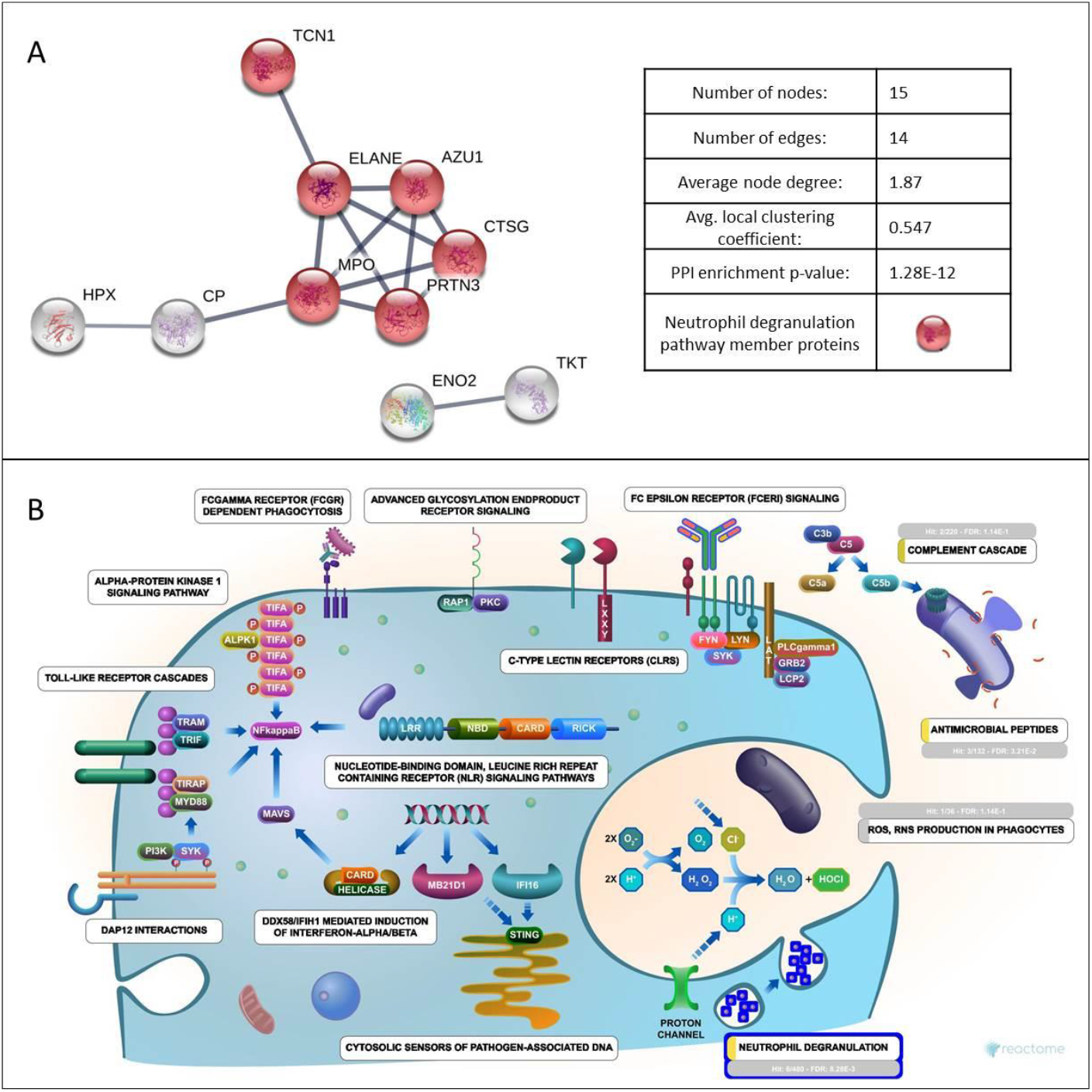
STRING and Reactome analysis of identified proteins. A-Protein-protein interaction network. B-Diagram of Initiate Immune System pathway (HSA-168249, FDR=2.3E-5) from Reactome, showing a significant enrichment for neutrophile degranulation pathway (HSA-6798695, FDR=2.01E-7) having the most hits.

## 4. Discussion

Neutrophils play a vital role in airway infections and may also define the disease outcome and carry out various functions in viral infections ranging from phagocytosis, degranulation to the generation of neutrophil extracellular traps (NETs) [3]. Neutrophils also interact with the immune cells and secreting cytokines participate in eliciting the antiviral response. Degranulation mechanism is part of the innate immune system which is necessary for the fight against infection and provides the neutrophils with the necessary tools. In COVID-19 patients, it was argued that the interlink between the innate and adaptive immune system may increase the severity and mortality of the disease [4]. Granules in neutrophils carry synthesized antimicrobial molecules and the content differs depending on the stage of maturation [5], depending on their contents they are named as azurophilic, gelatinase granules, specific, and secretory vesicles [6].

In SARS-CoV-2 patients’ naso-oropharyngeal samples, we have identified azurophilic granule (AG) proteins like Myeloperoxidase (MPO), elastase (ELANE), cathepsin G (CTSG), azurocidin 1 (AZU1) and proteinase 3 (PRTN3) to be highly overexpressed. Azurophilic neutrophils undergo limited exocytosis when activated [7] and their primary role is believed to be killing and degradation of engulfed microbes in the phagolysosome [8]. Although neutrophil degranulation having a positive impact on clearance of SARS-CoV-2 from the nasopharynx, in the inflammation microenvironment the process is broken and may lead to tissue damage by the secreted molecules from the granules. naso-oropharyngeal samples of COVID-19 patients demonstrate an increased level of both PRTN3 and MPO. PRTN3, a serine protease enzyme that is involved in granulocyte differentiation is expressed in the neutrophil granulocytes [9]. Promyelocyte proliferation is inhibited by PR3 expression that leads to neutrophil differentiation [10]. PRTN3 has various catalytic functions like cleavage of extracellular matrix and antiinflammatory proteins and activation of cytokines. MPO which is also expressed in neutrophil granulocytes can generate chlorine oxidants to ill infection agents [11]. Anti-neutrophil cytoplasmic antibodies (ANCA) autoantibodies specifically target azurophil proteins like PRTN3 and MPO and their role in numerous autoimmune disorders are discussed but primary focuses is systemic vasculitis where blood vessels are destroyed via inflammation [10]. ANCA with cytoplasmic staining pattern against PRTN3 (c-ANCA) is seen in granulomatosis with polyangiitis (GPA) whereas perinuclear staining pattern against MPO (p-ANCA) is seen in eosinophilic granulomatosis with polyangiitis(EGPA), microscopic polyangiitis (MPA), and pauci-immune idiopathic crescentic glomerulonephritis [12]. Vascular endothelium in GPA patients are damaged by neutrophils that actively participate in autoimmune response [13]. Vasculitis risk in patients increases with PRTN3 positive neutrophils [14]. PRTN3 is expressed on the surface of the neutrophils and the ANCAs that target PRTN3 causes pro-inflammatory effects [9]. The increased PRTN3 and MPO production in the neutrophils of COVID-19 patients are the target for anti-neutrophil cytoplasmic antibodies. Increased PRTN3 was shown to have a negative effect on the resolution of inflammation that causes immune system deregulation [15]. In a mouse study, ANCAs that target MPO were injected through IV and it triggered extra capillary glomerulonephritis [16]. Neutrophil apoptosis is necessary in the resolution of inflammation [17]. The resolution of inflammation and tissue repair processes are aided by the cytokines released from macrophages during phagocytosis of the neutrophils [10]. The dysregulation in the macrophage orchestrated phagocytosis and neutrophil apoptosis mechanisms leads to inflammation [18]. Patients with ANCA originated vasculitis are more prone to apoptosis as a result of ANCA activation [19]. PRTN3 is reported as a signal that sustains the inflammatory microenvironment [9]. The overexpression of PRTN3 observed in COVID-19 patients may impair the resolution of inflammation during neutrophil apoptosis. It is argued that the recruited neutrophils to the site of inflammation get activated and the release of uncontrolled mediators may cause tissue damage. The inflammation seen in vasculitis is converted to necrotizing vasculitis via the release of toxic mediators due to early neutrophil apoptosis and also by decreased level of apoptotic neutrophil clearance by macrophages [10]. PRTN3 was also reported to interfere with complement protein C1-q mediated clearance of Apoptotic Cells [20].

NETosis is a type of programmed cell death where neutrophil extracellular traps are formed [21]. NETs were identified in 2004 and they are often overlooked as drivers of severe pathogenic inflammation [22]. NETs have pathogen killing properties and include strands of DNA wrapped with histones and are enriched with neutrophil proteins like MPO and ELANE [23]. PRTN3, ELANE, and CTSG are coined as neutrophil serine proteases (NSP) and have tissue degrading and microbial killing effects [24]. NSP’s active role in neutrophil-associated lung inflammatory and tissue-destructive diseases has been reported [25]. Increased expressions of PRTN3, ELANE, and CTSG was reported in COPD patients [26]. In a mouse model study ELANE or PRTN3 was introduced to the trachea which caused tissue destruction and enlargement in airspace [27]. ELANA expression was also implicated in the impairment of host defense resulting in a decrease in mucociliary clearance of bacteria and also pathogens phagocytosis [28]. CD2, CD4, and CD8 can be cleaved on the surface of T-cells by ELANE and CTSG that dysregulate T-cell Function [29]. It was reported that patients that lack alpha-1 antitrypsin (α1-Pi) which s the physiological inhibitor of PRTN3 and ELANE carries a high risk of developing emphysema [30]. Azurophil granules also carry Cathepsin D which was reported to be up-regulated in the pulmonary macrophages in a mouse model of cigarette smoking [31]. Severe COPD cases exhibited MPO positive cells as a signal of neutrophil activation [32]. On the other hand in a mouse model of influenza it was shown that the inflammation damage was reduced by the absence of MPO [33]. IL-32, a proinflammatory cytokine with four isoforms, cleavage by PRTN3 propagates cytokine activity and triggers IL-1beta, TNF-alpha, IL-6, and chemokines [34]. It was argued that the targeted inhibition of PR3 or silencing of IL-32 by an inactive form of PRTN3 may halt the IL-32 driven immune dysregulation [34]. NSPs like ELANE, CTSG, and PRTN3 can cleave pro-IL-1beta to bioactive IL-1beta [35]. Caspase-1/Interleukin-1 converting enzyme (ICE) cleaves proteins like precursors of the inflammatory cytokines interleukin 1β and interleukin 18 into their mature biologically active forms [36] and CTSG regulates Caspase-1 in this pathway [37]. ICE has an active role in cell immunity as an initiator of inflammatory response so once activated it triggers the formation of active IL-1beta which is secreted from the cell that induces inflammation in the neighboring cells [38]. It was recently reported that infection in COVID-19 patients with acute respiratory syndrome showed release of the pro-inflammatory cytokines like IL-1beta and IL-6 [39]. Through the available literature, we can see that the up-regulation of various proteins observed in the naso-oropharyngeal swab samples of COVID-19 patients is tightly interconnected with the immune response.

## 5. Conclusions

The alterations of various proteins in SARS-CoV-2 infected patients’ naso-oropharyngeal samples depict the molecular changes that govern the host antiviral defense system. The available literature for many respiratory diseases are very helpful in linking altered protein expressions to viral pathogenesis. Obtained data provided us an important view of SARS-CoV-2 molecular changes on the protein level in the infection site. Statistically significant protein alterations of PRTN3, MPO, ELANE, CTSG, AZU1, and TCN1 dysregulation is important in the early phases of infection and may be targets for anti-SARS-CoV-2 therapeutics. Further research may show a link between the level of these proteins with disease severity and may be used as prognostic markers. Modulating the dysregulated proteins like PRTN3 or MPO may promote an anti-inflammatory response to alleviate SARS-CoV-2 symptoms. Also targeting NETs to dampen the out-of-control host response as a treatment may increase the survival rate by reducing the number of patients who require mechanical ventilation in ICU. We posit here that excess NETs may elicit the severe multi-organ consequences of COVID-19 via their known effects on tissues and the immune, vascular, and coagulation systems. Targeting NSPs and NETs in COVID-19 patients should therefore be considered by the biomedical community.

## Data Availability

All supplementary material will be provided with the manuscript.

## Funding

No specific funding was received from profit or non-profit organizations.

## Credit authorship contribution statement

All authors have contributed to the study design and execution of the methodology. Sample collection, data acquisition, and data processing was done by Ahmet Tarik Baykal, Mete Bora Tuzuner, Emel Akgun, Betul Sahin, Mustafa Serteser, Ibrahim Unsal, Meltem Kilercik, Canan Kulah and Hacer Nur Cakiroglu. All authors have contributed and approved the manuscript.

## Conflict of Interest/Disclosure Statement

The authors have declared no conflict of interest.

## Abbreviations

CE: collision energy
DIA: data-independent acquisition
FA: formic acid
UPLC: Ultra performance liquid chromatography
UPX: Universal protein extraction kit.

## Appendix A. Supplementary data

Supplementary Table 1. Protein quantification data

Supplementary Table 2. Protein identification data

## Notes

### Competing Interest Statement

The authors have declared no competing interest.

## References

[1] Busra Gurel, Mehmet Cansev, Cansu Koc, Busra Ocalan, Aysen Cakir, Sami Aydin, Nevzat Kahveci, Ismail H Ulus, Betul Sahin, Merve Karayel Basar, Ahmet Tarik Baykal (2019). Proteomics Analysis of CA1 Region of the Hippocampus in Pre-, Progression and Pathological Stages in a Mouse Model of the Alzheimer’s Disease. Curr Alz Res. http://doi:10.2174/1567205016666190730155926

[2] Moseley MA, Hughes CJ, Juvvadi PR, Soderblom EJ, Lennon S, Perkins SR, Thompson JW, Steinbach WJ, Geromanos SJ, Wildgoose J, Langridge JI, Richardson K, Vissers JPC (2018). Scanning quadrupole data‐independent acquisition, part A: qualitative and quantitative characterization. J Proteome Res. https://doi.org/10.1021/acs.jproteome.7b00464

[3] Jeremy V. Camp and Colleen B. Jonsson (2017). A Role for Neutrophils in Viral Respiratory Disease. Front Immunol. https://doi:0.3389/fimmu.2017.00550

[4] Sean Quan Du, Weiming Yuan (2020). Mathematical Modeling of Interaction between Innate and Adaptive Immune Responses in COVID-19 and Implications for Viral Pathogenesis. J of Med Vir. https://doi.org/10.1002/jmv.25866

[5] Kathleen D. Metzler, Christian Goosmann, Aleksandra Lubojemska, Arturo Zychlinsky, and Venizelos Papayannopoulos (2014). A Myeloperoxidase-Containing Complex Regulates Neutrophil Elastase Release and Actin Dynamics during NETosis. Cell Rep. https://doi:10.1016/j.celrep.2014.06.044

[6] Mahalakshmi Ramadass and Sergio D Catz (2016). Molecular mechanisms regulating secretory organelles and endosomes in neutrophils and their implications for inflammation. Immunol Rev. https://doi:10.1111/imr.12452

[7] Sengeløv H, Kjeldsen L, Borregaard N (1993). Control of exocytosis in early neutrophil activation. J Immunol. 15;150(4):1535–43

[8] K A Joiner, T Ganz, J Albert, D Rotrosen (1989). The opsonizing ligand on Salmonella typhimurium influences incorporation of specific, but not azurophil, granule constituents into neutrophil phagosomes. J Cell Biol. https://doi.org/10.1083/jcb.109.6.2771

[9] Kai Kessenbrock, Leopold Fröhlich, Michael Sixt, Tim Lämmermann, Heiko Pfister, Andrew Bateman, Azzaq Belaaouaj, Johannes Ring, Markus Ollert, Reinhard Fässler, and Dieter E. Jenne (2008). Proteinase 3 and neutrophil elastase enhance inflammation in mice by inactivating antiinflammatory progranulin. J Clin Invest. https://doi:10.1172/JCI34694

[10] V’eronique Witko-Sarsat Nathalie Thieblemont (2017). Granulomatosis with polyangiitis (Wegener granulomatosis): a proteinase-3 driven disease? Joint Bone Spine. http://dx.doi.org/doi:10.1016/j.jbspin.2017.05.004

[11] Seymour J. Klebanoff, Anthony J. Kettle, Henry Rosen, Christine C. Winterbourn, and William M. Nausee (2013). Myeloperoxidase: a front-line defender against phagocytosed microorganisms. J Leukoc Biol. http://doi:10.1189/jlb.0712349

[12] Johannes Schulte-Pelkum, Antonella Radice, Gary L. Norman, Marcos López Hoyos, Gabriella Lakos, Carol Buchner, Lucile Musset, Makoto Miyara, Laura Stinton, and Michael Mahler (2014). Novel Clinical and Diagnostic Aspects of Antineutrophil Cytoplasmic Antibodies. J Immun Res. http://dx.doi.org/10.1155/2014/185416

[13] Lise Halbwachs and Philippe Lesavr (2012). Endothelium-Neutrophil Interactions in ANCA-Associated Diseases. J Am Soc Nephrol. http://doi:10.1681/ASN.2012020119

[14] S Ohlsson, J Wieslander, and M Segelmark (2003). Increased circulating levels of proteinase 3 in patients with anti-neutrophilic cytoplasmic autoantibodies-associated systemic vasculitis in remission. Clin Exp Immunol. http://doi:10.1046/j.1365-2249.2003.02083.x

[15] Witko-Sarsat V, Reuter N, Mouthon L (2010). Interaction of proteinase 3 with its associated partners: implications in the pathogenesis of Wegener’s granulomatosis. Current opinion in rheumatology. 22:1–7.

[16] Lani Shochet, Stephen Holdsworth and A. Richard Kitching (2020). Animal Models of ANCA Associated Vasculitis. Front. Immunol. https://doi.org/10.3389/fimmu.2020.00525

[17] Katherine R. Martin, Delphine Ohayona, and Véronique Witko-Sarsa (2015). Promoting apoptosis of neutrophils and phagocytosis by macrophages: novel strategies in the resolution of inflammation. BIOMEDICAL INTELLIGENCE. https://doi.org/10.4414/smw.2015.14056

[18] Yumiko Oishi and Ichiro Manabe (2016). Macrophages in age-related chronic inflammatory diseases. NPJ Aging Mech Dis. https://doi:10.1038/npjamd.2016.18

[19] Mohamed Abdgawad, Åsa Pettersson, Lena Gunnarsson, Anders A. Bengtsson, Pierre Geborek, Lars Nilsson, Mårten Segelmark and Thomas Hellmark (2012). Decreased Neutrophil Apoptosis in Quiescent ANCA-Associated Systemic Vasculitis. PLoS One. https://doi:10.1371/journal.pone.0032439

[20] Pascale Tacnet-Delorme, Julie Gabillet, Simon Chatfield, Nathalie Thieblemont, Philippe Frachet and Véronique Witko-Sarsat (2018). Proteinase 3 interferes With c1q-Mediated clearance of apoptotic cells. Fron. Immun. https://doi:10.3389/fimmu.2018.00818

[21] Tobias A. Fuchs, Ulrike Abed, Christian Goosmann, Robert Hurwitz, Ilka Schulze, Volker Wahn, Yvette Weinrauch, Volker Brinkmann, Arturo Zychlinsky (2007). Novel cell death program leads to neutrophil extracellular traps. JCB. https://doi.org/10.1083/jcb.200606027

[22] Brinkmann V, Reichard U, Goosmann C, Fauler B, Uhlemann Y, Weiss DS, Weinrauch Y, Zychlinsky A. (2004). Neutrophil extracellular traps kill bacteria. Science. https://DOI:10.1126/science.1092385

[23] Yanbao Yu, Keehwan Kwon, Tamara Tsitrin, Shiferaw Bekele, Patricia Sikorski, Karen E. Nelson, and Rembert Pieper (2017). Characterization of Early-Phase Neutrophil Extracellular Traps in Urinary Tract Infections. PLoS Pathog. https://doi:10.1371/journal.ppat.1006151

[24] Brice Korkmaz, Marshall S. Horwitz, Dieter E. Jenne, and Francis Gauthier (2010). Neutrophil Elastase, Proteinase 3, and Cathepsin G as Therapeutic Targets in Human Diseases. Pharmacol Rev. https://doi:10.1124/pr.110.002733

[25] Kalupov T, Brillard-Bourdet M, Dadé S, Serrano H, Wartelle J, Guyot N, Juliano L, Moreau T, Belaaouaj A, Gauthier F (2009). Structural characterization of mouse neutrophil serine proteases and identification of their substrate specificities: relevance to mouse models of human inflammatory diseases. J Biol Chem. https://doi:10.1074/jbc.M109.042903

[26] Almansa R, Socias L, Sanchez-Garcia M, Martín-Loeches I, del Olmo M, Andaluz-Ojeda D, Bobillo F, Rico L, Herrero A, Roig V, San-Jose CA, Rosich S, Barbado J, Disdier C, de Lejarazu RO, Gallegos MC, Fernandez V, Bermejo-Martin JF (2012). Critical COPD respiratory illness is linked to increased transcriptomic activity of neutrophil proteases genes. BMC Res Notes. https://doi:10.1186/1756-0500-5-401

[27] Lafuma C, Frisdal E, Harf A, Robert L, Hornebeck W (1991). Prevention of leucocyte elastase-induced emphysema in mice by heparin fragments. Eur Respir J. 4(8):1004–9.

[28] Raquel Almansa, Lorenzo Socias, Monica Sanchez-Garcia, Ignacio Martín-Loeches, Milagros del Olmo, David Andaluz-Ojeda, Felipe Bobillo, Lucia Rico, Agueda Herrero, Vicente Roig, C Alicia San-Jose, Sara Rosich, Julia Barbado,8 Carlos Disdier, Raúl Ortiz de Lejarazu, Maria C Gallegos, Victoria Fernandez, and Jesus F Bermejo-Martin (2012). Critical COPD respiratory illness is linked to increased transcriptomic activity of neutrophil proteases genes. BMC Res Notes. https://doi:10.1186/1756-0500-5-401

[29] G Döring, F Frank, C Boudier, S Herbert, B Fleischer and G Bellon (1995). Cleavage of lymphocyte surface antigens CD2, CD4, and CD8 by polymorphonuclear leukocyte elastase and cathepsin G in patients with cystic fibrosis. J Immunol. 154 (9) 4842–4850

[30] Mario Cazzola, Daiana Stolz, Paola Rogliani, Maria Gabriella Matera (2020). α1-Antitrypsin deficiency and chronic respiratory disorders. European Respiratory Review. https://DOI:10.1183/16000617.0073-2019

[31] Ken R BrackeDidier CataldoTania MaesTania MaesShow all 8 authorsRomain A Pauwels (2005). Matrix Metalloproteinase-12 and Cathepsin D Expression in Pulmonary Macrophages and Dendritic Cells of Cigarette Smoke-Exposed Mice. International Archives of Allergy and Immunology. https://DOI:10.1159/000088439

[32] Di Stefano A, Caramori G, Ricciardolo FL, Capelli A, Adcock IM, Donner CF (2004). Cellular and molecular mechanisms in chronic obstructive pulmonary disease: an overview. Clin Exp Allergy. https://doi:10.1111/j.1365-2222.2004.02030.x

[33] Sugamata R, Dobashi H, Nagao T, Yamamoto K, Nakajima N, Sato Y, Aratani Y, Oshima M, Sata T, Kobayashi K, Kawachi S, Nakayama T, Suzuki K (2012). Contribution of neutrophil-derived myeloperoxidase in the early phase of fulminant acute respiratory distress syndrome induced by influenza virus infection. Microbiol Immunol. https://doi:10.1111/j.1348-0421.2011.00424.x.

[34] Daniela Novick, Menachem Rubinstein, Tania Azam, Aharon Rabinkov, Charles A. Dinarello, and Soo-Hyun Kim (2006). Proteinase 3 is an IL-32 binding protein. PNAS. https://doi.org/10.1073/pnas.0511206103

[35] Monica Guma, Lisa Ronacher, Ru Liu-Bryan, Shinji Takai, Michael Karin and Maripat Corr, M (2009). Caspase-1 Independent IL-1β Activation in Neutrophil Dependent Inflammation. Arthritis Rheum. https://doi:10.1002/art.24959

[36] Hella LukschMichael RomanowskiMichael RomanowskiOsvaldo CharaOsvaldo CharaShow all 26 authorsAngela Rösen-WolffAngela Rösen-Wolff (2013). Naturally Occurring Genetic Variants of Human Caspase-1 Differ Considerably in Structure and the Ability to Activate Interleukin-1β. Human Mutation. https://10.1002/humu.22169

[37] Lalitha Srinivasan, Sarah Ahlbrand, and Volker Briken (2014). Interaction of Mycobacterium tuberculosis with Host Cell Death Pathways. Cold Spring Harb Perspect Med. https://doi:10.1101/cshperspect.a022459

[38] Marcus A Lawson, Robert H McCusker & Keith W Kelley (2013). Interleukin-1 beta converting enzyme is necessary for development of depression-like behavior following intracerebroventricular administration of lipopolysaccharide to mice. Journal of Neuroinflammation. https://doi:10.1186/1742-2094-10-54

[39] Conti P, Ronconi G, Caraffa A, Gallenga CE, Ross R, Frydas I and Kritas SK (2020). Induction of pro-imflammatory cytokines (IL-1 and IL-6) and lung inflammation by Coronavirus-19 (CoV-19 or SARS-CoV-2): anti-inflammatory strategies. J Biol Regul Homeost Agents. http://doi:10.23812/CONTI-E

